# Subset-based method for cross-tissue transcriptome-wide association studies improves power and interpretability

**DOI:** 10.1101/2023.01.11.23284454

**Authors:** Xinyu Guo, Nilanjan Chatterjee, Diptavo Dutta

**Affiliations:** Department of Quantitative and Computational Biology, University of Southern California; Department of Biostatistics, Bloomberg School of Public Health, Johns Hopkins University; Department of Oncology, School of Medicine, Johns Hopkins University; Integrative Tumor Epidemiology Branch, Division of Cancer Epidemiology & Genetics, National Cancer Institute

## Abstract

Integrating results from genome-wide association studies (GWAS) and studies of molecular phenotypes like gene expressions, can improve our understanding of the biological functions of trait-associated variants, and can help prioritize candidate genes for downstream analysis. Using reference expression quantitative trait loci (eQTL) studies, several methods have been proposed to identify significant gene-trait associations, primarily based on gene expression imputation. Further, to increase the statistical power by leveraging substantial eQTL sharing across tissues, meta-analysis methods aggregating such gene-based test results across multiple tissues or contexts have been developed as well. However, most existing meta-analysis methods have limited power to identify associations when the gene has weaker associations in only a few tissues and cannot identify the subset of tissues in which the gene is “activated” in. For this, we developed a novel cross-tissue subset-based meta-analysis (CSTWAS) method which improves power under such scenarios and can extract the set of potentially “active” tissues. To improve applicability, CSTWAS uses only GWAS summary statistics and pre-computed correlation matrices to identify a subset of tissues that have the maximal evidence of gene-trait association. We further developed an adaptive monte-carlo procedure with the generalized Pareto distribution (GPD) to accurately estimate highly significant p-values for the test statistics. Through numerical simulations, we found that CSTWAS can maintain a well-calibrated type-I error rate, improves power especially when there is a small number of “active” tissues for a gene-trait association and identifies an accurate “active” tissue-set. By analyzing several GWAS summary statistics of three complex traits and diseases, we demonstrated that CSTWAS could identify novel biological meaningful signals while providing an interpretation of disease etiology by extracting a set of potentially “active” tissues.

## Introduction

Genome-wide association studies (GWAS) have been immensely successful in identifying hundreds and thousands of genetic variants associated with a spectrum of diseases and traits^1,2^. Despite this success, the majority of genetic variants identified by GWAS are in the noncoding region^3–5^; hence, their functions are often challenging to interpret. Several studies have shown that the variants identified through GWAS are enriched in gene regulatory elements^6,7^. Further, these variants have substantial overlap with those associated with different molecular phenotypes, including gene expression levels^8^, protein levels^9^, metabolites^10^, and others^11^, indicating that they might have a regulatory role in the disease etiology. Thus, large-scale studies integrating such molecular data with genetic studies to identify potentially associated genes and intermediate molecular phenotypes have become increasingly important.

Transcriptomic studies that identify expression quantitative trait loci (eQTLs) meaning, variants associated with the expression levels of a gene in a particular tissue, have become one of the standard approaches to understand the regulatory role of variants. By integrating eQTL information of variants with the results of GWAS, a number of statistical tools have been developed to identify potential target genes through which genetic associations may be mediated^12–15^. Given the lack of availability of data on outcome, genetic variants, and gene expression simultaneously on the same individuals, the major body of methodological research in this domain has adopted a two-step approach. First, a genetically regulated expression (GReX) score is constructed for a given gene, based on the local genetic variants (usually within 500kb ∼ 1Mb) using a reference gene expression study. Pre-computed models for constructing GReX using reference transcriptomic studies like GTEx^16^, TCGA^17^, and others^18–20^ are publicly available. Subsequently, we test the association of this constructed GReX with the outcome, for significance. If significant, we can infer that the expression levels of the gene have a significant effect on the outcome, and this test can be conducted for each gene across the transcriptome. In the post-GWAS era, such transcriptome-wide association studies (TWAS) have emerged as an effective downstream GWAS analysis^21^ to identify trait-associated candidate genes. Many methods have been developed for conducting such TWAS using individual-level as well as summary-level data, like PrediXcan (for individual-level GWAS data)^12^, S-PrediXcan^13^, and FUSION (for summary-level GWAS data)^14^.

The overall power of such expression-imputation methods depends on building an accurate predictive GReX from the reference data^22^. However, most gene expression studies till date have relatively small sample sizes^16,17^. Further, the usual approach of TWAS is to perform an agnostic scan across all or multiple available tissues to identify tissue-gene pairs significantly associated with the outcome, which can increase the multiple testing burden. One potential way to improve the power of such tests is to aggregate or meta-analyze results across multiple tissues or contexts. Although gene expression depends on various factors, like tissue type, cell type, developmental stages, and others^23,24^, there are substantial overlap between eQTLs across multiple related tissues^16^, which can be effectively exploited to share information through meta-analysis. Therefore, to increase the power of TWAS results, existing approaches either leverage information across multiple tissues or meta-analyze results from multiple tissues, such as UTMOST^25^, MultiXcan^15^ and FUSION-OMNIBUS^14^.

However, even though these standard meta-analysis methods have proved to be effective in increasing the power to identify gene-trait associations, there are some important limitations. Firstly, these methods tend to demonstrate lower power in situations where the causal gene is activated in only a small number of tissues. Furthermore, it remains important to identify the tissues in which a trait-associated gene is “active”. Due to aggregation across multiple tissues, the current methods lack information on the set of potentially activated tissues for a gene-trait association, thus failing to provide effective interpretability.

Here, we developed a cross-tissue subset-based meta-analysis (CSTWAS) method that aims to increase power over existing methods while maintaining proper type-I error control and improves interpretation by extracting a subset of tissues in which the gene is potentially activated. Similar approaches have been utilized previously in the context of single variant meta-analysis^26^, multiple phenotypes^27^, gene-sets^28^, and gene-environment interactions^29^. CSTWAS, being a meta-analysis, can leverage association summary statistics across multiple tissues or contexts, and increase statistical power by aggregating weaker associations. Compared to standard meta-analysis approaches, the major advantage of CSTWAS is that it can identify a potential “active” subset of tissues or contexts, which helps in interpreting the association of the gene with the outcome. Further, through simulations, we show that CSTWAS can improve statistical power over standard approaches, especially when the number of active tissues for a gene is relatively low and maintains comparable power otherwise. CSTWAS only needs summary-level GWAS data and pre-computed covariance matrices (only need to be computed once for each transcriptomic reference panel) and hence can be performed with publicly available data. Through analysis of publicly available GWAS summary statistics for three distinct traits and diseases, we show that CSTWAS can identify important associations missed by standard methods as well as provide effective interpretation.

## Methods

CSTWAS (Cross-tissue Subset-based Transcriptome Wide Association Study) is a subset-based meta-analysis of individual gene-based test results across different tissues resulting in a p-value corresponding to the null hypothesis that there is no association of the gene with the outcome across any of the tissues. Additionally, it outputs a set of potential active tissues for each gene with aggregated evidence of association, which can be interpreted as the set of active tissues. Starting from GWAS summary statistics, CSTWAS can be implemented in the following steps (**Figure 1**):

1. **Perform gene-based tests with summary-level GWAS data across multiple tissues:** Using GWAS summary statistics, perform standard gene-based expression imputation tests of associations (e.g., S-PrediXcan, FUSION, or others) for a given gene across different available tissues. In simulations and data analysis, we have used p-values from FUSION test with GTEx v7 reference panel.
2. **Construct the test statistic for the gene by selecting a set of potential active tissues:** Next, we convert the p-values of the gene across different tissues to z-values, which quantifies the evidence of association on a gaussian scale. Let *p*_1_, *p*_2_, …, *p*_*T*_ be the FUSION p-values for specific gene expression across T tissues, we first convert these p-values to z-values *z*_1_, *z*_2_, …, *z*_*T*_ as:

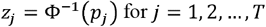

Where, Φ^−1^ is the inverse standard normal cumulative distribution function. Suppose *z*_*j*_ is the converted z-value for a specific gene expression in tissue *j, H* is the entire set of tissues (such as all 48 tissues in GTEx v7), *B* is a non-empty subset of tissues in *H*, and |*B*| is the number of tissues in subset *B*, we construct a test statistic that maximizes the average association among all possible subsets of tissues.

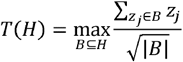

*T* : the number of tissues *H* : the entire set of tissues *B* : a subset of tissues |*B*|: the number of tissues in subset *B* Since there are 2^*T*^ − 1 non-empty possible subset *B*, calculation of this test statistics is computationally expensive, especially with a large number of reference tissues. To select the subset of tissues (*B*) more efficiently, we simplified the enumeration process by:
  - Sort all z-values in decreasing order:

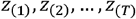
  - Calculate the test statistics, starting with the largest z-value:

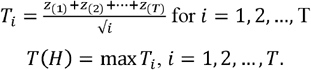
3. **Calculating p-values by a stepwise approach:** The tissue-specific association test statistics (*z*_*i*_) are correlated due to the correlation among GReX of multiple related tissues. So, it is not straightforward to derive the analytical null distribution of the CSTWAS test statistic. Instead, we adopt an efficient simulation-based approach. We first estimated the null covariance for 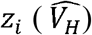 across T tissues:
  - For a specific gene across all tissues, ran gene-based test (we used FUSION here) analysis with a simulated null phenotype and obtained the converted z-values *z*_1_, *z*_2_, …, *z*_*T*_
  - Repeated the previous step *K* (=1000) times, getting a *K* × *N* matrix for the gene.
  - Calculated the covariance matrix 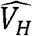 across tissue-specific association statistics (*z*_*i*_)

**Figure 1:**
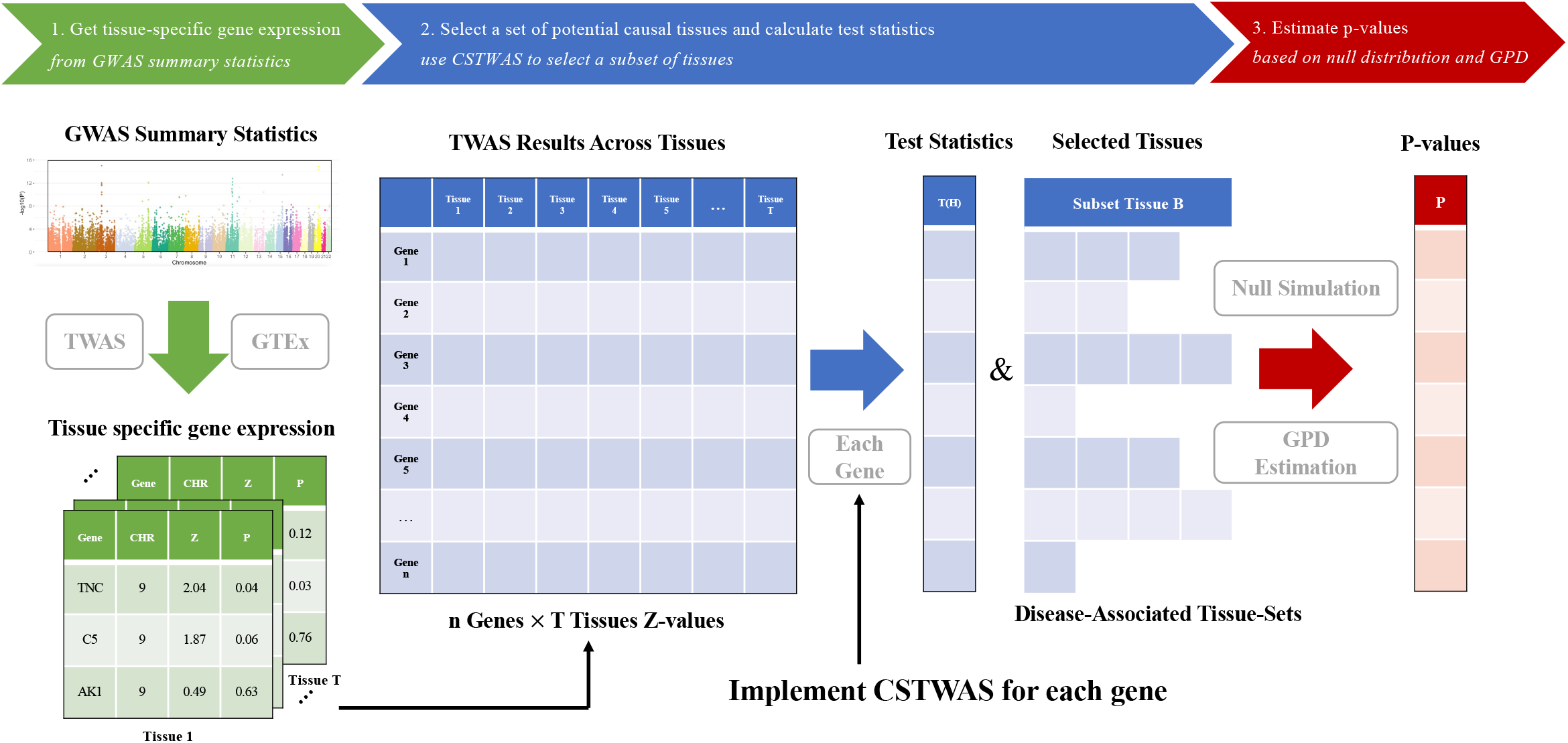
Overview of CSTWAS Methods. 1. Construct tissue-specific GReX levels from GWAS summary-level statistics with GTEx as the reference panel. Here, many tools can be utilized, such as S-Predixcan and FUSION. 2. Aggregate gene expression information across tissues in a matrix. Then, for each gene expression, run CSTWAS to select a set of active tissues, aggregating the evidence of the associations between GReX and complex trait. 3. Estimate p-values for each CSTWAS statistic with null simulation and GPD estimation.

Given the estimated null covariance for 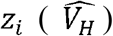, we assume that under the null hypothesis that the gene has no association with the outcome for any tissues, the tissue-specific association test statistics matrix Z follows a multivariant normal distribution.

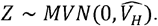

Subsequently, we estimate the p-value through a Monte Carlo (or simulation) procedure:

- Generated a null gene expression z-value vector across tissues from the multivariate normal distribution.
- Calculated the test statistics *T*(*H*)_*null*_ from simulated z-values and sorted decreasingly.
- Repeated previous two steps M times and estimated the p-value by

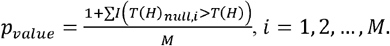

It is to be noted that the precision of the p-value estimation is constrained by the number of monte-carlo iterations (*M* in this case). However, if there is strong evidence to support the identified set of potential active tissues, the p-value would be very small. Hence accurately estimating the low p-value by simulation would be computationally expensive. Therefore, we utilized the generalized Pareto distribution (GPD)-based tail estimation method^30^ to estimate p-values less than 10^−6^. Overall, we fit the GPD to the right tail of the simulated test statistics and estimate the p-value by inverting the distribution function of the GPD. More specifically, we first model the right tail of test statistics by GPD. The GPD has the density function

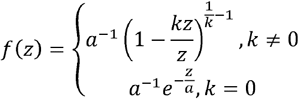

and cumulative distribution function

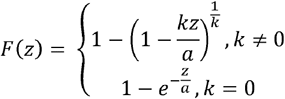

Where, *a* is the scale parameter and *k* is the shape parameter. Also, 0 ≤ *z* < ∞ for *k* ≤ 0 and 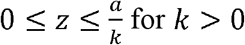. We use the MLE approach^31^ to estimate the GPD distribution with the right tail of test statistics. The scale parameter *a* can be estimated by

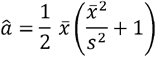

and the shape parameter *k* can be estimated by

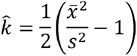

Here, 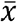 and *s*^2^ are the mean and the standard deviation of simulated test statistics. With the estimated parameters for GPD, we can construct the p-value as

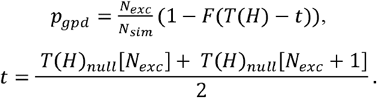

Here, *N*_*exc*_ is the exceedances threshold (usually use 250), *N*_*sim*_ is the total number of the simulated test statistics, *F* (.) is the GPD cumulative distribution, *T*(*H*) is the test statistics, and *t* is the threshold value of simulated statistics. In this case, 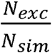 controls that the estimation only focuses on the right tail of simulated test statistics.

## Results

### Simulations

We used numerical simulations to evaluate the type-I error rate and power of CSTWAS in comparison to standard methods of analysis. To reflect realistic LD patterns, we used genotypes for *N* unrelated individuals in the cis-region of a given gene *G* from UK Biobank. Using this, first, we estimate the GReX for *G* across *T* different tissues. This can be done using the publicly available reference models precomputed from transcriptomic studies. In particular, we use the reference FUSION models in GTEx v7 for this. Further, we normalize the GReX such that for a given tissue, the GReX across *N* individuals have mean = 0 and variance = 1. We denote the normalized estimated GReX for individual *i* at the *t*^*th*^ tissues as *r*_*i*(*t*)_, *i* = 1, 2, …, *N* and *t* = 1, 2, …, *T*. We next simulated a continuous phenotype for the *i*^*th*^ individual under a normal error model as:

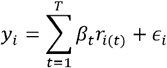

Where *β*_*t*_ denotes the effect of the GReX for the *t*^*th*^ tissue and *ϵ*_*i*_ is the normal error term. Subsequently, we conducted the GWAS using the individual-level genotype and simulated phenotype data and further conducted the FUSION test using the GWAS summary statistics to extract the gene-level p-values across *T* tissues for which pre-computed reference models of gene *G* were available in GTEx v7. Using these outputs from this gene-level test, we perform the cross-tissue subset-based TWAS. Across all the simulation scenarios, we set the sample size *N* to be unrelated White British individuals. We performed the simulations for two different genes: *APOE* (*T* = 24) and *ABO* (*T* = 45).

### Type-I error

To evaluate the type-I error rate, we set each *β*_*t*_ = 0. Hence the phenotype *y*_*i*_ is generated independently of the genotypes from a univariate standard normal distribution. For both the genes, the results (**Table 1**) show that the type-I error rate for the cross-tissue subset-based test is relatively well calibrated across different levels (1 × 10^−4^, 1 × 10^−5^) and at the exome-wide threshold (2.5 × 10^−6^). This indicates that the chances of false discovery through this method are controlled at the desired level.

**Table 1:** Type-1 error rates for the CSTWAS for genes *APOE* and *ABO*. Empirical type-1 error rates across different levels, expressed as the proportion of corresponding levels.

| Gene | Level | Type-1 error/Level |
| --- | --- | --- |
| *APOE* | $1 \times 10^{-04}$ | 0.98 |
| | $1 \times 10^{-05}$ | 0.99 |
| | $2.5 \times 10^{-06}$ | 1.01 |
| *ABO* | $1 \times 10^{-04}$ | 0.99 |
| | $1 \times 10^{-05}$ | 1.01 |
| | $2.5 \times 10^{-06}$ | 1.02 |

### Power

We compared the power of the CSTWAS test to several standard approaches: (a) Bonferroni corrected minimum p-value across all available tissues for a gene, which we call minP, (b) FUSION-Omnibus, which performs a multiple degree-of-freedom fixed effects meta-analysis of FUSION results across tissues for the gene and (c) UTMOST, which performs cross-tissue association test using a generalized Berk-Jones statistic^32^ and performs well in moderately sparse scenarios. As mentioned above, we focus on two different genes: *APOE* and *ABO*, which are significantly expressed in *T* = 24 and *T* = 45 tissues respectively. To evaluate the power of CSTWAS in comparison to the other methods, we randomly designated *T*_*a*_ tissues within the *T* tissues to be active, meaning the tissues for which the gene has non-zero effects on the phenotype. For these tissues, we generated the corresponding *β*_*t*_ from a normal distribution such that *β*_*t*_ ∼ *N* (0, *v*), where *v* is a variance parameter that was selected such that, the total variance explained by the gene in active tissues (*h*^2^) remained at a desired level (usually between 1-5%; **Figure 2**). We simulated a continuous phenotype using the above Gaussian error model, performed the FUSION across the *T* tissues and recorded the corresponding p-values. Subsequently, we applied CSTWAS, FUSION-Omnibus, and UTMOST to the results and calculated the empirical power as the proportion of simulation iterations that identified the gene to be significant at an exome-wide threshold of 2.5 × 10^−6^. To reflect a spectrum of possible association scenarios, we studied the empirical power across various values of and *T*_*a*_ and *h*^2^.

**Figure 2:**
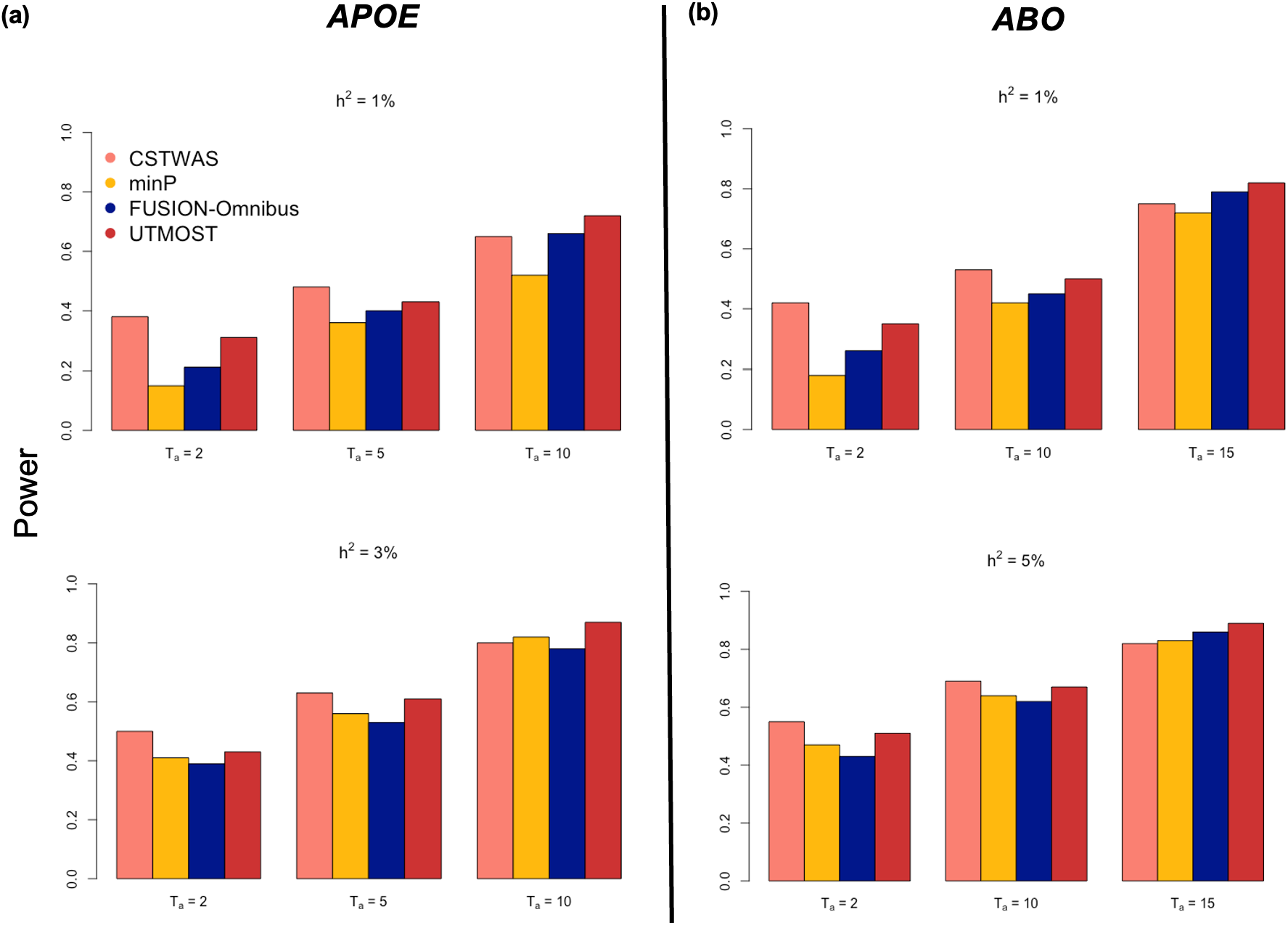
Simulation Results. Empirical power for CSTWAS. Simulation results with (a) *APOE* gene and (b) *ABO* gene. Estimated empirical power for CSTWAS compared to other methods for different values of h^2^ and number of active tissues (*T*_*a*_). See Simulation for more details.

The results (**Figure 2**) show that no test is uniformly most powerful across all the scenarios. We find that for both genes, CSTWAS is substantially more powerful under a sparse scenario, meaning a smaller number of active tissues (*T*_*a*_). UTMOST has a slightly lower power compared to CSTWAS, in such scenarios, with the difference in power decreasing with increasing number of active tissues. With a larger number for *T*_*a*_, FUSION-Omnibus and minP also, have a comparable or better performance than CSTWAS. This is potentially because, for smaller *T*_*a*_ with weaker signals, an increased number of null values for a larger number of tissues dilutes the signal and hence lowers the power. However, our method, being subset-based, can separate an optimal subset of tissues with potentially weaker signals. The pattern remains consistent across different *h*^2^ values, although the difference in power at low *T*_*a*_ is reduced for higher *h*^2^ which corresponds to the scenario where the gene has strong signal in each tissue.

Since, one principal motivation and advantage of CSTWAS is identifying potentially active tissues for a gene-trait association, we further inspected the sensitivity and specificity of the selected subset of tissues. Sensitivity is defined as the proportion of active tissues correctly identified by CSTWAS in the selected subset, and specificity is defined as the proportion of inactive genes correctly identified by CSTWAS. Because no other methods currently identify the active tissues, we compared the performance of CSTWAS to minP by defining the significant tissues (Bonferroni corrected p-value < 2.5*×*10^−6^) as active. We performed this numerical experiment with *ABO* gene only and set the total variance explained by the gene in active tissues (*h*^2^) to be 3% throughout. Across a spectrum different number of active tissues, CSTWAS maintained a higher sensitivity and specificity, with both the values decreasing, with increasing *T*_*a*_ as expected (**Supplementary Figure 1**). However, for low *T*_*a*_, we found a substantial difference in specificity between the methods, indicating that the minP might select some correlated tissues as well, while CSTWAS accounting for the cross-tissue correlation pattern, selects the active set with high accuracy.

Simulation results highlight the utility of CSTWAS compared to the standard methods. Especially in a scenario, when the gene is activated weakly in only a few tissues, CSTWAS has substantially higher power to identify gene-trait associations which might not be adequately captured by FUSION-Omnibus or minP, while the performance of UTMOST is comparable. However, CSTWAS provides an additional biological interpretation by extracting the potential set of tissues where the gene is active and might be associated with the outcome.

#### Association analysis of three traits and diseases using CSTWAS

We applied CSTWAS to publicly available GWAS summary statistics for three different representative diseases and trait outcomes. For that, we constructed a null covariance matrix () for each gene using the FUSION reference models provided for GTEx v7. Further, we compared the results and genes identified by CSTWAS to those identified by FUSION in each tissue adjusting for multiple comparisons, which we call tissue-specific TWAS.

##### Bipolar Disorder (BD)

Bipolar disorder is a chronic psychiatric disorder that affects about 60 million people worldwide. Although primarily neurological, recently evidence has emerged that the genetic etiology of BD has complex involvement of multiple distinct tissues other than brain^33^. To investigate that, we applied CSTWAS to the bipolar disorder GWAS summary statistics provided by the Psychiatric Genetics Consortium (PGC), containing 29,764 cases and 169,118 controls^34^. Using CSTWAS, we identified 205 genes that are significantly associated with BD at an α level of 2.5 × 10^−6^ which is the exome-wide significance level using the Bonferroni correction for 20,000 genes (**Figure 3**). Among these, 55 associations are novel in that they are identified through CSTWAS alone and are not significant in the tissue-specific TWAS results. Meaning, these genes did not have a strong or significant association with BD in any of the individual tissues and can only identified by aggregating weaker associations through CSTWAS. Several of the novel genes identified have evidence in existing literature to be associated with BD and other neuropsychiatric processes (**Table 2**). For example, *ZNF280D*, a protein-coding gene, is closely related to its family member *ZNF804A*, which has been identified as an important factor in bipolar disorder development^35^; *SULT1A3*, another protein-coding gene, has negative effects on sulfation and the impaired sulfation has been proved to have associations with many neurological diseases, such as bipolar disease^36^. Since such gene-based results primarily identify genetic loci, where multiple highly correlated genes can reside, we additionally clumped (See **Supplementary** for details) the associations based on genomic positions to identify the unique loci or genomic regions identified by each test. CSTWAS identifies 82 unique and significant loci while 67 significant loci are identified by tissue-specific TWAS. Out of these, 57 were also identified by CSTWAS, indicating a high concordance between the methods along with several novel discoveries through CSTWAS (**Supplementary Figure 2**).

**Table 2:** Significant genes associated with Bipolar disorder (BD) identified by CSTWAS. Several example genes associated to BD, their CSTWAS p-value, minimum TWAS p-value across all available tissues in GTEx v7 and extracted set of potentially active tissues are shown. See **Supplementary Table 1** for full results. Tissues related to central nervous system are highlighted in bold.

| Gene | CSTWAS Tissue-set | CSTWAS P-value | Min TWAS P-value |
| --- | --- | --- | --- |
| *FAM65A* | **Brain_Caudate_basal_ganglia**, **Brain_Hypothalamus**, Ovary, Heart_Left_Ventricle, Thyroid, **Brain_Cerebellar_Hemisphere**, **Brain_Anterior_cingulate_cortex_BA24**, Colon_Sigmoid | $8.62 \times 10^{-07}$ | $4.43 \times 10^{-06}$ |
| *SULT1A3* | **Brain_Cerebellar_Hemisphere**, Heart_Atrial_Appendage, Muscle_Skeletal, Thyroid, Artery_Coronary, Uterus | $2.32 \times 10^{-06}$ | $5.95 \times 10^{-06}$ |
| *RARRES3* | Skin_Not_Sun_Exposed_Suprapubic, **Brain_Caudate_basal_ganglia**, **Brain_Cerebellum**, Colon_Transverse, **Brain_Substantia_nigra**, Artery_Tibial, Colon_Sigmoid, Stomach, Liver | $8.75 \times 10^{-07}$ | $5.97 \times 10^{-06}$ |
| *AC105339.1* | **Brain_Caudate_basal_ganglia**, **Brain_Amygdala**, Spleen, **Brain_Nucleus_accumbens_basal_ganglia**, **Brain_Anterior_cingulate_cortex_BA24**, Pituitary, Skin_Sun_Exposed_Lower_leg | $1.24 \times 10^{-06}$ | $8.85 \times 10^{-05}$ |
| *MEI4* | Heart_Atrial_Appendage, Brain_Cortex, Esophagus_Muscularis, Testis, **Brain_Nucleus_accumbens_basal_ganglia**, Pituitary, Artery_Aorta, Brain_Cerebellar_Hemisphere, **Brain_Anterior_cingulate_cortex_BA24**, Skin_Not_Sun_Exposed_Suprapubic, Artery_Coronary, **Brain_Putamen_basal_ganglia**, **Brain_Hippocampus**, **Brain_Caudate_basal_ganglia**, Heart_Left_Ventricle, **Brain_Cerebellum**, **Brain_Amygdala**, Esophagus_Gastroesophageal_Junction | $1.77 \times 10^{-08}$ | $2.98 \times 10^{-05}$ |
| *ZNF280D* | Breast_Mammary_Tissue, **Brain_Cerebellar_Hemisphere**, Vagina, Cells_Transformed_fibroblasts, Pancreas, Whole_Blood, **Nerve_Tibial**, Skin_Not_Sun_Exposed_Suprapubic, Thyroid, Heart_Left_Ventricle, Lung, **Brain_Frontal_Cortex_BA9** | $4.80 \times 10^{-07}$ | $6.89 \times 10^{-06}$ |
| *NTM* | Heart_Left_Ventricle, Testis, Artery_Coronary, **Brain_Anterior_cingulate_cortex_BA24**, Breast_Mammary_Tissue, Esophagus_Mucosa, **Brain_Spinal_cord_cervical_c.1**, Cells_Transformed_fibroblasts, Skin_Sun_Exposed_Lower_leg, Esophagus_Gastroesophageal_Junction, **Nerve_Tibial**, Lung | $2.72 \times 10^{-10}$ | $5.96 \times 10^{-11}$ |

**Table 3:** Significant genes associated with Breast Cancer (BC) identified by CSTWAS. Several example genes associated to BC, their CSTWAS p-value, minimum TWAS p-value across all available tissues in GTEx v7 and extracted set of potentially active tissues are shown. See **Supplementary Table 2** for full results. Tissues related to breast, female-specific tissues, and tissues which are known to be involved in BC are highlighted in bold.

| Gene | CSTWAS Tissue-set | CSTWAS P-value | Min TWAS P-value |
| --- | --- | --- | --- |
| *WDR37* | **Vagina**, Adipose_Visceral_Omentum, **Adipose_Subcutaneous**, Skin_Sun_Exposed_Lower_leg, Nerve_Tibial, **Ovary**, Colon_Transverse, Brain_Spinal_cord_cervical_c.1, Small_Intestine_Terminal_Ileum, Stomach, Esophagus_Muscularis, Lung, Skin_Not_Sun_Exposed_Suprapubic, **Breast_Mammary_Tissue**, **Thyroid**, Prostate, Artery_Tibial, Brain_Caudate_basal_ganglia, Brain_Hippocampus, Esophagus_Gastroesophageal_Junction, Heart_Left_Ventricle, Brain_Nucleus_accumbens_basal_ganglia, Uterus, Artery_Aorta, **Whole_Blood**, Artery_Coronary | $6.72 \times 10^{-07}$ | $2.70 \times 10^{-05}$ |
| *FAT4* | Pancreas, Esophagus_Gastroesophageal_Junction, Brain_Caudate_basal_ganglia, Muscle_Skeletal, Prostate, **Uterus**, Skin_Sun_Exposed_Lower_leg, Brain_Spinal_cord_cervical_c.1, Spleen | $1.79 \times 10^{-06}$ | $8.75 \times 10^{-06}$ |
| *FGF7* | Nerve_Tibial, Heart_Left_Ventricle, Esophagus_Muscularis, Esophagus_Gastroesophageal_Junction, Muscle_Skeletal, Heart_Atrial_Appendage, Spleen, Brain_Hippocampus, Artery_Tibial, **Adipose_Subcutaneous**, **Thyroid** | $1.82 \times 10^{-07}$ | $2.82 \times 10^{-06}$ |
| *ORC2* | Colon_Sigmoid, Brain_Substantia_nigra, **Cells_EBV.transformed_lymphocytes**, **Breast_Mammary_Tissue**, **Uterus**, **Ovary**, **Whole_Blood**, Heart_Left_Ventricle, Esophagus_Muscularis | $6.41 \times 10^{-07}$ | $1.05 \times 10^{-07}$ |
| *CALCRL* | **Uterus**, Brain_Nucleus_accumbens_basal_ganglia, Pituitary, **Breast_Mammary_Tissue**, Adipose_Subcutaneous, **Ovary**, Artery_Aorta, Artery_Tibial, Muscle_Skeletal, **Whole_Blood**, **Thyroid**, Nerve_Tibial, Artery_Coronary, Esophagus_Gastroesophageal_Junction, Skin_Sun_Exposed_Lower_leg | $4.12 \times 10^{-06}$ | $7.83 \times 10^{-06}$ |

**Table 4:** Significant genes associated with Serum Urate Level (SUL). Several example genes associated to SUL, their CSTWAS p-value, minimum TWAS p-value across all available tissues in GTEx v7 and extracted set of potentially active tissues are shown. See **Supplementary Table 3** for full results. Liver and lower alimentary tissues are highlighted in bold.

| Gene | CSTWAS Tissue-set | CSTWAS P-value | Min TWAS P-value |
| --- | --- | --- | --- |
| *LRRC16A* | Ovary, Heart_Atrial_Appendage, Heart_Left_Ventricle, Muscle_Skeletal, Breast_Mammary_Tissue, **Colon_Transverse**, Adrenal_Gland, Brain_Caudate_basal_ganglia, Prostate, Skin_Not_Sun_Exposed_Suprapubic, Thyroid, Brain_Spinal_cord_cervical_c.1 | $2.39 \times 10^{-10}$ | $2.26 \times 10^{-41}$ |
| *ZGPAT* | Adipose_Subcutaneous, Brain_Cerebellar_Hemisphere, Artery_Aorta, Heart_Left_Ventricle, Skin_Not_Sun_Exposed_Suprapubic, Brain_Cortex, Cells_Transformed_fibroblasts, Whole_Blood, Muscle_Skeletal, **Small_Intestine_Terminal_Ileum, Liver**, Adipose_Visceral_Omentum, Skin_Sun_Exposed_Lower_leg | $1.08 \times 10^{-08}$ | $1.06 \times 10^{-05}$ |
| *ASGR1* | **Small_Intestine_Terminal_Ileum**, Uterus, Artery_Tibial, Esophagus_Gastroesophageal_Junction, Brain_Cerebellar_Hemisphere, Brain_Cerebellum, Adipose_Visceral_Omentum | $4.53 \times 10^{-07}$ | $4.44 \times 10^{-05}$ |
| *RP11-794G24.1* | Artery_Coronary, Vagina, Lung, Brain_Substantia_nigra, Ovary, **Colon_Sigmoid**, Brain_Hippocampus, **Small_Intestine_Terminal_Ileum**, Prostate, Minor_Salivary_Gland, Muscle_Skeletal, Heart_Left_Ventricle | $2.34 \times 10^{-06}$ | $5.65 \times 10^{-05}$ |
| *PDE8A* | Lung, Brain_Frontal_Cortex_BA9, Adipose_Subcutaneous, Cells_EBV.transformed_lymphocytes, **Small_Intestine_Terminal_Ileum, Liver**, Esophagus_Mucosa, Adipose_Visceral_Omentum, Colon_Transverse, Breast_Mammary_Tissue, Spleen, Brain_Amygdala, Brain_Cortex, Adrenal_Gland, Esophagus_Muscularis, Skin_Not_Sun_Exposed_Suprapubic, Brain_Hypothalamus, Artery_Aorta, Nerve_Tibial, Ovary | $1.12 \times 10^{-06}$ | $8.85 \times 10^{-05}$ |
| *PPP1R16B* | Breast_Mammary_Tissue, Brain_Cerebellum, Artery_Aorta, Esophagus_Gastroesophageal_Junction, Brain_Cerebellar_Hemisphere, Testis, Brain_Substantia_nigra, Artery_Tibial, Prostate, **Liver**, Colon_Transverse, Adipose_Visceral_Omentum, Nerve_Tibial, **Small_Intestine_Terminal_Ileum**, Adrenal_Gland | $1.36 \times 10^{-06}$ | $3.64 \times 10^{-05}$ |

**Figure 3:**
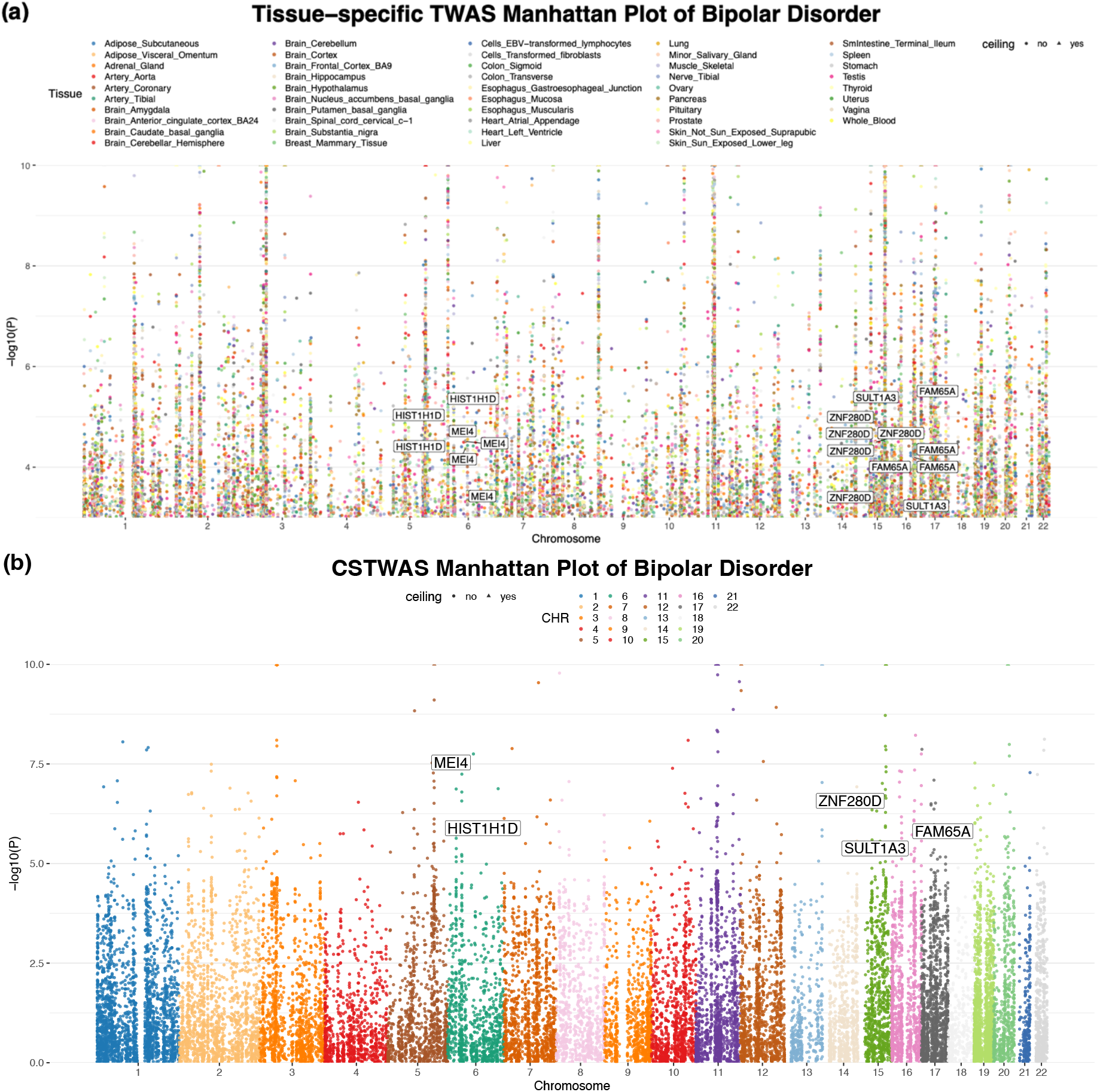
Results for tissue-specific TWAS across multiple tissues and CSTWAS of bipolar disorder (BD). (a) P-values for TWAS results in tissues reported in GTEx v7 using FUSION. Each dot represents the TWAS -log10(p-value) for a tissue-specific GReX association of bipolar disorder (BD) colored by tissues. Highly significant p-values (< 1 × 10^−10^: the ceiling cutoff) were shown by 1 × 10^−10^ (represented by triangles). P-values > 1 × 10^−03^ are hidden for ease of viewing. (b) P-values for CSTWAS. Each dot represents the p-value for integrated GReX associations of BD across tissues colored by chromosomes. P-values < 1 × 10^−10^ were shown by 1 × 10^−10^. Five example genes, *MEI4, HIST1H1D, ZNF280D, SULT1A3*, and *FAM65A*, are annotated in both plots, which do not have significant associations (P-values > 2.5 × 10^−6^) in any of the tissues but are significant (P-values < 2.5 × 10^−6^) using CSTWAS.

In addition to increased power, CSTWAS can also provide important insights into the identified associations by extracting active tissues (**Table 2** and **Supplementary Table 1**). For example, *NTM*, a neurotrimin protein-coding gene, is identified to be associated with BD (p-value = 2.72 × 10^−10^ ; See **Link Availability**). By considering each tissue independently, the tissue-specific TWAS only identified significant expression in Heart Left Ventricle (p-value = 5.96 × 10^−11^) which might not be readily interpretable in the context of neurological processes. However, the subset-based approach successfully detected the weaker associations in Brain Anterior cingulate cortex BA24 (tissue-specific p-value = 4.45 × 10^−4^), Brain Spinal cord cervical c.1 (tissue-specific p-value = 5.63 × 10^−3^). and Nerve Tibial (tissue-specific p-value = 4.65 × 10^−2^) all of which are relevant in context of BD. These weaker associations may be lower power to be detected due to the smaller reference panel size of these tissues (N = 126 to 532). Thus, CSTWAS provides a novel interpretation of the association of *NTM* with BD by extracting the potential active tissue set, which includes several weaker signals not identified in tissue-specific analysis. It is to be noted that for several of the top significant genes, alongside CNS tissues, other tissues are selected. This can be caused due to many factors, including high eQTL sharing between tissues and pleiotropy.

##### Breast Cancer (BC)

According to CDC, breast cancer is the second most common cancer among women in the U.S. Evidence has been shown that some BC development factors might not only be restricted to breast tissue but can span multiple cell types and organs^37^. To identify BC-related candidate genes and corresponding sets of active tissues, we applied CSTWAS to the breast cancer GWAS summary statistics, containing 17,881 cases and 410,350 controls^38^. Using CSTWAS, we identified 37 genes that are significantly associated with BC at an exome-wide threshold of 2.5 × 10^−6^ (**Figure 4**). Among these associations, 11 novel associations are identified as significant in CSTWAS and are not significant in the tissue-specific TWAS results, meaning they did not have any strong association across the different tissues. For example, CSTWAS identifies *FAT4* (p-value = 1.79 × 10^−6^), a protein-coding gene, which is identified to be a tumor suppressor in triple-negative breast cancer^39^ and is repressed in several cancers due to promoter hypermethylation; *FGF7* (p-value = 1.82 × 10^−7^), which increases breast cancer cell proliferation and migration in vitro, is also identified to be associated^40^. There are 25 significant loci (clumped as before) identified by CSTWAS and 20 significant loci identified by tissue-specific TWAS. Among these identified significant loci, 14 are overlapped between the two approaches, indicating CSTWAS can recover most of result from tissue-specific TWAS with some novel findings (**Supplementary Figure 3**).

**Figure 4:**
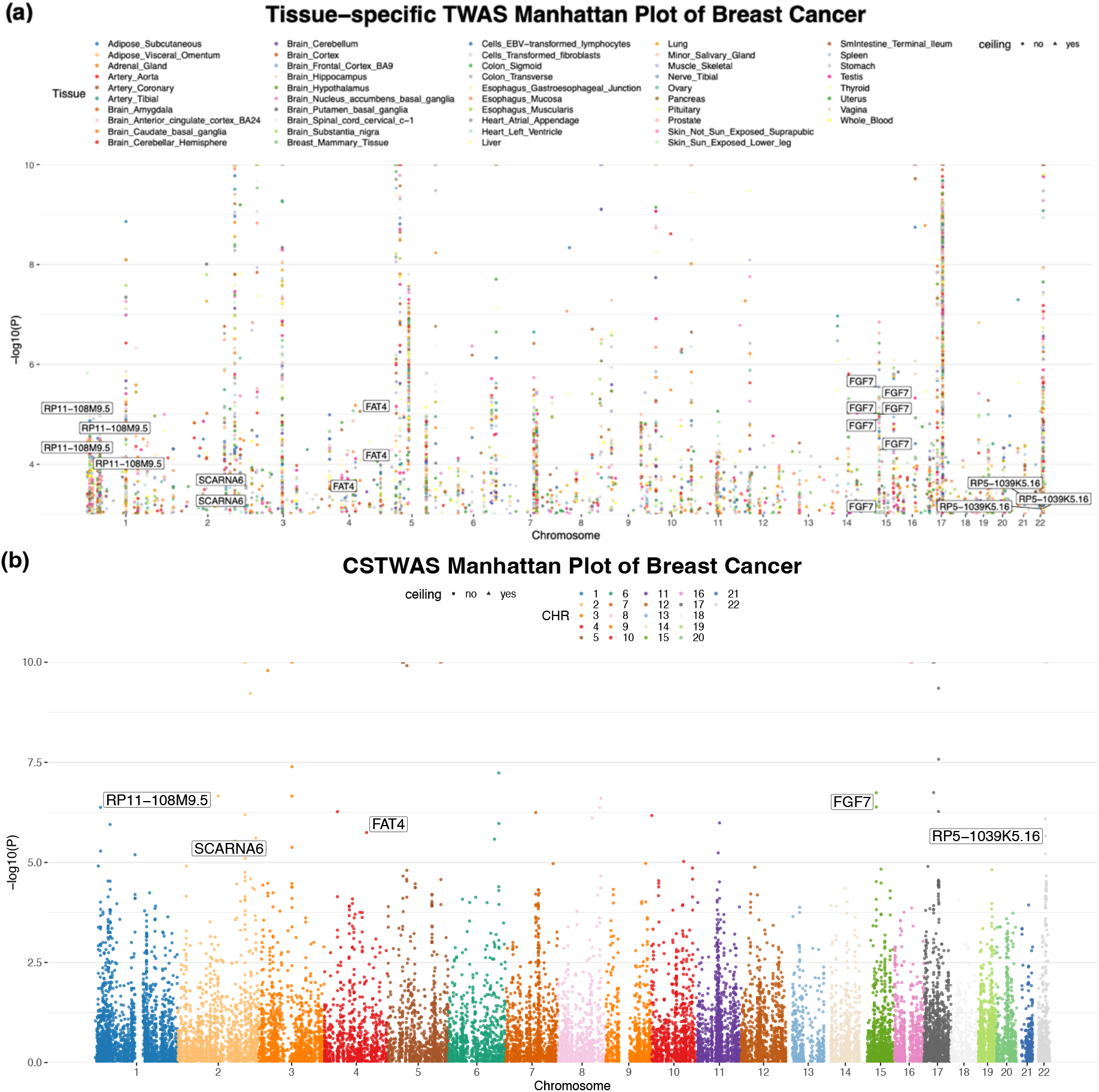
Results for tissue-specific TWAS across multiple tissues and CSTWAS of breast cancer (BC). (a) P-values for TWAS results in multiple tissues. Each dot represents the TWAS p-value for a tissue-specific gene-GReX association of breast cancer (BC). Different tissues are plotted in different colors. Here, P-values < 1 × 10^−10^ (the ceiling cutoff) were shown by 1 × 10^−10^ (represented by triangles). And P-values > 1 × 10^−3^ are hidden for simplicity. (b) P-values for CSTWAS. Each dot represents the p-value for integrated GReX associations of breast cancer (BC) across tissues. P-values < 1 × 10^−10^ were shown by 1 × 10^−10^. Five genes, *RP11-108M9*.*5, SCARNA6, FAT4, FGF7*, and *RP5-1039K5*.*16*, are annotated in both plots, which do not have significant associations (P-values > 2.5 × 10^−6^) in any of the tissues but are significant (P-values < 2.5 × 10^−6^) using CSTWAS.

Moreover, several genes have weaker associations, which CSTWAS can successfully identify but are not identified through tissue-specific TWAS. For example, CSTWAS identified *ORC2* (p-value = 6.41 × 10^−7^), a protein-coding gene, that has been shown to have abnormal behavior in many types of cancers, including breast cancer^41^. Tissue-specific TWAS only identified significant expression in Colon Sigmoid (p-value = 1.05 × 10^−7^). This result might not be readily interpretable for breast cancer. However, CSTWAS successfully detected the weaker signals—gene expression in Breast Mammary Tissue (tissue-specific p-value = 2.04 × 10^−7^), Ovary (tissue-specific p-value = 7.81 × 10^−3^) and Whole Blood (tissue-specific p-value = 3.55 × 10^−2^), which are all relevant tissues for BC. Thus CSTWAS, beyond improving power to identify associations, can additionally identify potential active tissue-set (p-value for overall association = 6.41 × 10^−7^), including the weaker signals which were not detected in the tissue-specific approach.

##### Serum Urate Level (SUL)

Human serum urate level is an indicator of uric acid production and excretion. A recent study has shown that SUL has a strong genetic component with heritability between 30% and 60% with the most relevant tissues for SUL being kidney and liver^42^. To identify potential genes and corresponding active tissues for SUL, we applied CSTWAS to the SUL GWAS summary statistics, containing 457,690 individuals^42^. In our analysis, we identified 243 genes that are significantly associated with SUL at an *α* level 2.5 × 10^−6^, which is the exome-wide significance level using the Bonferroni correction for 20,000 genes (**Figure 5**). Among these, 44 associations are novel that they are identified through CSTWAS alone and are not significant in the tissue-specific TWAS results. As before, we clumped the associations to identify the unique loci highlighted by each test. CSTWAS identifies 81 significant loci while 71 significant GReX associations identified by tissue-specific TWAS. Out of these, 65 were also identified by CSTWAS indicating high overlap with several novel discoveries (**Supplementary Figure 4**).

**Figure 5:**
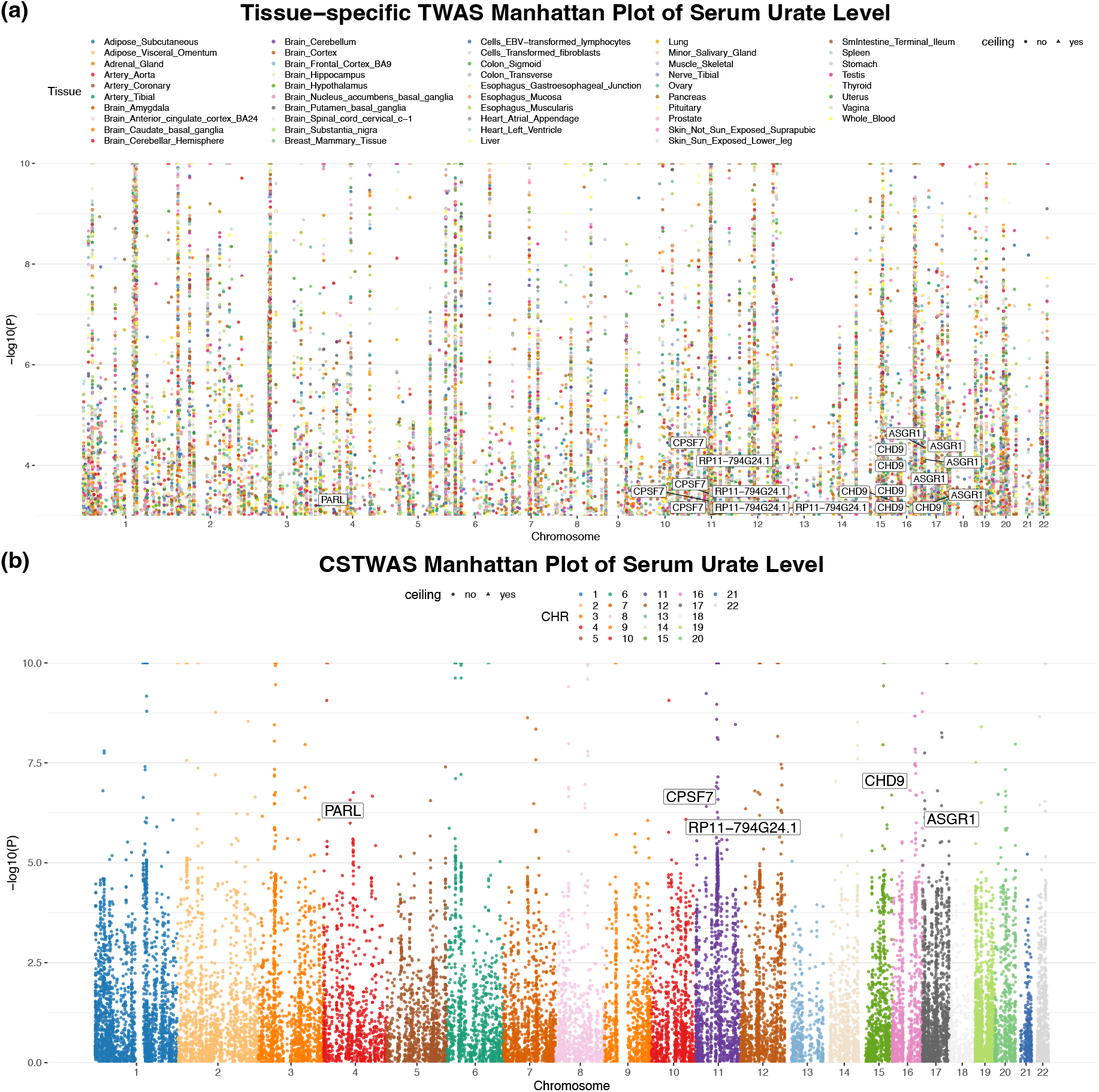
Results for tissue-specific TWAS across multiple tissues and CSTWAS of human serum urate levels (SUL). (a) P-values for TWAS results in multiple tissues. Each dot represents the TWAS p-value for a tissue-specific GReX association of human serum urate level (SUL). Different tissues are plotted in different colors. Here, P-values < 1 × 10^−10^ (the ceiling cutoff) were shown by 1 × 10^−10^ (represented by triangles). And P-values > 1 × 10^−3^ are hidden for simplicity. (b) P-values for CSTWAS. Each dot represents the p-value for integrated GReX associations of human serum urate levels (SUL) across tissues. P-values < 1 × 10^−10^ were shown by 1×10^−10^. Five genes, *PARL, CPSF7, RP11-794G24*.*1, CHD9*, and *ASGR1*, are annotated in both plots, which do not have significant associations (P-values > 2.5 × 10^−6^) in any of the tissues but are significant (P-values < 2.5 × 10^−6^) using CSTWAS.

We further demonstrate that, CSTWAS can also provide important insights into the identified associations by extracting potentially active tissues. For example, *LRRC16A*, a protein-coding gene, has been shown to be associated with SUL^43^. Tissue-specific TWAS only identified significant gene expression in Heart Atrial Appendage (p-value = 3.25 × 10^−12^), Heart Left Ventricle (p-value = 5.89 × 10^−12^), Muscle Skeletal (p-value = 2.93 × 10^−9^), Ovary (p-value = 2.26 × 10^−41^) which might be difficult to interpret since most of these tissues are not directly involved in the serum urate processes. However, CSTWAS, on the other hand, successfully detected additional weaker signals—*LRRC16A* gene expression in Colon Transverse (tissue-specific p-value = 5.25 × 10^−7^) and Thyroid (tissue-specific p-value = 5.98 × 10^−4^), which have been shown to be relevant to SUL processes. Thus, CSTWAS identified the potential active tissue set (p-value for overall association = 2.39 × 10^−10^), including the weaker signals which were not identified in the tissue-specific approach, possibly due to the smaller reference panel size of these tissues (N = 305 to 574) and provided an improved interpretation to the association of *LRRC16A*.

## Discussions

Here we developed a subset-based cross-tissue meta-analysis method, CSTWAS, that integrates gene-based results across multiple tissues to identify associations of a gene with an outcome, and additionally identifies a potential set of tissues in which the gene is activated. Through simulations, we showed CSTWAS maintains desired type-I error rate and can outperform standard methods in several scenarios, especially where the gene has a weaker association in a small number of tissues. Also, through analysis of existing GWAS summary statistics of three traits and diseases, we demonstrated that CSTWAS could successfully detect weaker signals as well as provide interpretations for corresponding results.

The gain of statistical power over standard methods (like UTMOST, FUSION Omnibus or tissue-specific TWAS) is one of the key advantages of CSTWAS. The standard meta-analysis-based approaches fail to identify gene-trait association in scenarios, where the association signature is present in only a few tissues, since the majority of the current methods aim to aggregate results across all the tissues instead of choosing the “active” tissues. In the presence of few weaker associations, the signal is diluted by many null associations. In contrast, CSTWAS can extract a subset of tissues that are maximally associated with the outcome and hence improve statistical power by considering only potential associations. Further, the simulation results show that the CSTWAS can control type-I errors at exome-wide level as well. However, when the gene is activated in many tissues, the UTMOST or FUSION-OMNIBUS approach would provide higher or comparable power by selecting substantive associations.

One of the most prominent advantages of CSTWAS is the subset-based approach provides an interpretation of disease etiology through a potential subset of active tissues. The resulting subset of tissues can be interpreted as the tissues where the gene is actively mediating disease-related signals. While standard meta-analysis methods provide only a p-value for the association of the gene with the outcome, our method additionally identifies the potential active subsets of tissues and contexts as well. In addition, unlike the standard subset-based method searches across all possible subsets, which is exponentially scaled as the number of elements increases, we used an order-based method to improve the computational efficiency, which has previously been proposed in the context of gene-set associations and gene-environment associations. CSTWAS can search the potential active tissue set with computational complexity *O(nlogn)*. Further, CSTWAS can be requires GWAS summary statistics as input only, using a correlation matrix which can be calculated using publicly available reference data. In our software implementation, we provide this precomputed correlation matrix that can be used with FUSION results.

However, CSTWAS also has some limitations. Since the method highly relies on the correlation structure of a gene, across tissues, it is difficult to distinguish between highly correlated tissues, such as brain tissues. Hence selection consistency needs to be further studied. Further, ancestry mismatches between GWAS and transcriptomic reference panels may lead to increased false discoveries and complicate interpretation. In addition, when constructing the test statistics, instead of enumerating through all possible subsets, our method uses ordered Z-statistics for computational efficiency, which loses the directions of effects. In the future, we plan to extend our method to efficiently incorporate both direction and magnitude of effects in the analysis.

Overall, CSTWAS provides a powerful approach to meta-analyze tissue-specific gene-based test results to identify active tissues for an associated gene. The simulations and analysis of GWAS summary statistics have demonstrated the statistical properties and computational efficiency of CSTWAS. However, meta-analyzing results across tissues is one of the many possible applications of our generalizable subset-based approach. Like expressions across tissues, the gene expressions are also highly correlated across multiple cell types, developmental stages, and contexts. With massive publicly available single-cell data as well as emerging cell-type-specific eQTL studies, the subset-based approach can be translated to identify cell types and stages where genes are actively associated with diseases. In the future, we will seek applications and adaptations of the CSTWAS to different multi-omics data, which will provide further insights into the complex genetic architecture of diseases.

## Data Availability

All data produced in the present work are contained in the manuscript

https://github.com/Thewhey-Brian/CSTWAS

## Link & Data Availability

Gene description (*NTM*): https://www.ncbi.nlm.nih.gov/gtr/genes/50863/791

Gene description (*IDI1*): https://www.genecards.org/cgi-bin/carddisp.pl?gene792=IDI1keywords=IDI1793

Bipolar Disorder GWAS Summary Statistics: https://doi.org/10.6084/m9.figshare.14671998

Serum Urate Level GWAS Summary Statistics: http://ckdgen.imbi.uni-freiburg.de

Breast Cancer GWAS Summary Statistics: https://www.ebi.ac.uk/gwas/studies/GCST90011804

GitHub for CSTWAS package: https://github.com/Thewhey-Brian/CSTWAS

