## Supplementary for "Subset-based method for cross-tissue transcriptome-wide association studies improves power and interpretability"

**Supplementary Notes**

*Gene-Clumping*: A particular genomic loci can contain multiple genes expressed in different sets of tissues. Gene-based tests like FUSION identifies tissue-specific association of a gene with the trait in a particular tissue. Hence, the results of FUSION would be guided by the expression of patterns of genes in different tissues in the locus. Primarily, the gene-based tests identify a significant locus associated with the trait or disease in a particular tissue. To identify unique loci identified, we adopted a gene clumping approach. For each tissue-specific association using FUSION, we created a clumped list of genes with TSS within 200Kb. We tagged each locus by this list nearby genes. If two loci had overlapping genes, they were merged to create a united list of loci. This produced a list of unique loci identified by tissue-specific FUSION. We similarly clumped the associations identified by CSTWAS to identify and compared them with the list obtained from tissue-specific FUSION results for overlap.

**Supplementary Figures**

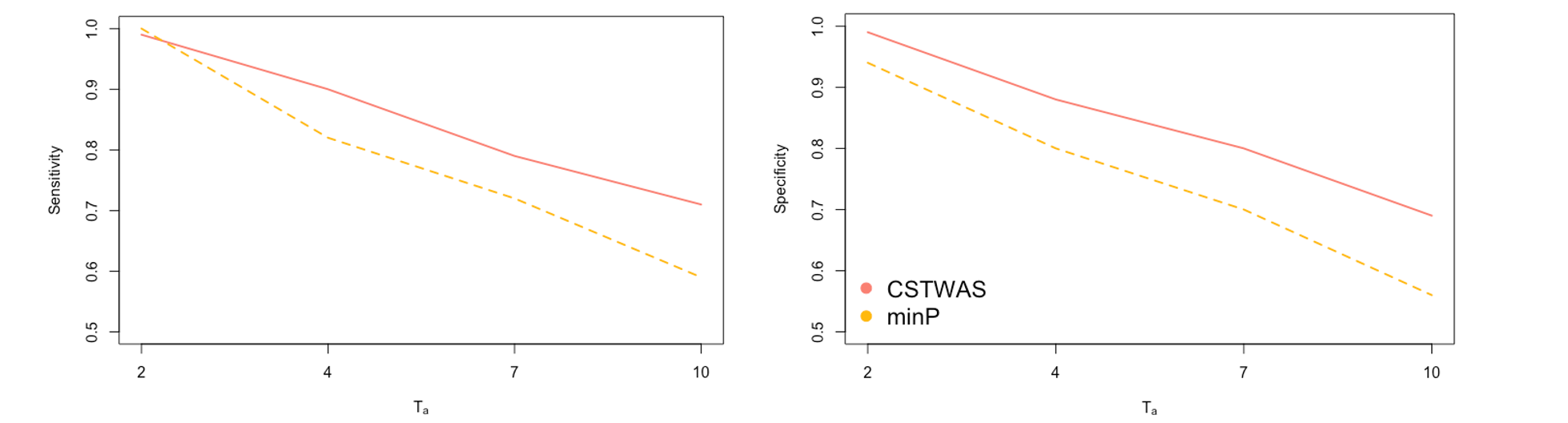

**Supplementary Figure 1: Sensitivity and specificity of CSTWAS.** The simulations were carried out for *ABO* (*T* = 45) gene only with $h^{2}=3\%$ across a spectrum of different number of active tissues (*T_a_*).

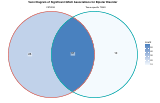

**Supplementary Figure 2**: Venn Diagram of significant GReX associations between CSTWAS (red border) and tissue-specific TWAS (blue border) for bipolar disorder (BD). The number of significant GReX associations in each category is labeled and color-coded by the intensity. The shared significant GReX associations are defined as their gene regions overlapping within a 200kb window.

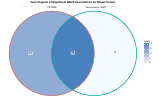

**Supplementary Figure 3**: Venn Diagram of significant GReX associations between CSTWAS (red border) and tissue-specific TWAS (blue border) for breast cancer (BC). The number of significant GReX associations in each category is labeled and color-coded by the intensity. The shared significant GReX associations are defined as their gene regions overlapping within a 200kb window.

**Supplementary Figure 4**: Venn Diagram of significant GReX associations between CSTWAS (red border) and tissue-specific TWAS (blue border) for serum urate level (SUL). The number of significant GReX associations in each category is labeled and color-coded by the intensity. The shared significant GReX associations are defined as their gene regions overlapping within a 200kb window.
